# Combined Bioinformatic and Splicing Analysis of Likely Benign Intronic and Synonymous Variants Reveals Evidence for Pathogenicity

**DOI:** 10.1101/2023.10.30.23297632

**Authors:** Owen R. Hirschi, Stephanie A. Felker, Surya P. Rednam, Kelly L. Vallance, D. Williams Parsons, Angshumoy Roy, Gregory M. Cooper, Sharon E. Plon

## Abstract

**Background:** Current clinical variant analysis pipelines focus on coding variants and intronic variants within 10-20 bases of an exon-intron boundary that may affect splicing. The impact of newer splicing prediction algorithms combined with *in vitro* splicing assays on rare variants currently considered Benign/Likely Benign (B/LB) is unknown.

**Methods:** Exome sequencing data from 576 pediatric cancer patients enrolled in the Texas KidsCanSeq study were filtered for intronic or synonymous variants absent from population databases, predicted to alter splicing via SpliceAI (>0.20), and scored as potentially deleterious by CADD (>10.0). Total cellular RNA was extracted from monocytes and RT-PCR products analyzed. Subsequently, rare synonymous or intronic B/LB variants in a subset of genes submitted to ClinVar were similarly evaluated. Variants predicted to lead to a frameshifted splicing product were functionally assessed using an *in vitro* splicing reporter assay in HEK-293T cells.

**Results:** KidsCanSeq exome data analysis revealed a rare, heterozygous, intronic variant (NM_177438.3(*DICER1*):c.574-26A>G) predicted by SpliceAI to result in gain of a secondary splice acceptor site. The proband had a personal and family history of pleuropulmonary blastoma consistent with *DICER1* syndrome but negative clinical sequencing reports. Proband RNA analysis revealed alternative *DICER1* transcripts including the SpliceAI-predicted transcript.

Similar bioinformatic analysis of synonymous or intronic B/LB variants (n=31,715) in ClinVar from 61 Mendelian disease genes yielded 18 variants, none of which could be scored by MaxEntScan. Eight of these variants were assessed (*DICER1* n=4, *CDH1* n=2, *PALB2* n=2) using *in vitro* splice reporter assay and demonstrated abnormal splice products (mean 66%; range 6% to 100%). Available phenotypic information from submitting laboratories demonstrated *DICER1* phenotypes in 2 families (1 variant) and breast cancer phenotypes for *PALB2* in 3 families (2 variants).

**Conclusions:** Our results demonstrate the power of newer predictive splicing algorithms to highlight rare variants previously considered B/LB in patients with features of hereditary conditions. Incorporation of SpliceAI annotation of existing variant data combined with either direct RNA analysis or *in vitro* assays has the potential to identify disease-associated variants in patients without a molecular diagnosis.

## Background

Identifying variants in clinically relevant genes that impact splicing is challenging yet essential for accurate genetic diagnosis for many diseases. Current variant classification guidelines from the American College of Medical Genetics and Genomics/Association for Molecular Pathology (ACMG/AMP) only assert that variants within 2 base pairs of a splice junction in a gene of interest are pathogenic candidates if loss-of-function of that gene is a known disease mechanism (1). Intronic variants farther away from these splice sites, however, may also affect splicing and be overlooked by these criteria. Such variants can result in the loss of the normal donor, acceptor, and branch point required for accurate mRNA splicing and instead result in creation of alternative splice products that alter gene function, potentially leading to loss-of-function. Variants predicted to be synonymous with regard to the protein sequence may also disrupt splicing; yet these variants are similarly deprioritized in clinical pipelines and may be penalized by commonly used predictive metrics like Combined Annotation Dependent Depletion (CADD) score, which incorporates coding consequence in its prediction of deleteriousness (2).

Intronic or synonymous variants affecting splicing can be rescued in genomic analysis with the use of bioinformatic tools such as SpliceAI, a deep neural network predictor of splice site activity in the pre-mRNA sequence (3). The tool uses genomic variation as input and outputs the position of the acceptor or donor site loss or gain as well as a predictive “delta score,” which is the maximum probability of splicing events affected by the variant within a user-determined window flanking the variant. SpliceAI is an improvement upon previous approaches, such as MaxEntScan, which detects splicing variants only if they disrupt canonical splicing motifs within 9bp on the donor (5’) splice site, and 23bp of the acceptor (3’) splice site, thus unable to analyze variants deeper into introns and exons (4).

Here, we demonstrate the added value of SpliceAI to identify rare variants previously classified as Likely Benign in *DICER1*, *CDH1*, and *PALB2.* These are all hereditary cancer genes with important clinical significance, and these variants result in aberrant splicing products, which create premature stop codons (PTC) in subsequent functional analysis of *in vitro* assays and patient RNA when available.

## Methods

### Texas KidsCanSeq Cohort

The Texas KidsCanSeq (KCS) study is a Clinical Sequencing Evidence-Generating (CSER) Consortium (5) study that recruited pediatric cancer probands under 18 years of age. The study was approved by Baylor College of Medicine institutional review board which served as the central IRB for all six participating sites. Probands and participating parents submitted blood or saliva samples for parallel clinical germline hereditary cancer panel and exome sequencing (6), with results reported back to the medical record. Consent included permission to perform subsequent research analyses with data shared with the CSER consortium (7).

### Cohort Variant Analysis and Variant Filtration

The germline exome variant call files (VCFs) from Texas KCS pediatric cancer probands were analyzed for variants within a list of 181 cancer predisposition genes (Additional File 1: Table S1). Variants were filtered to retain those with a total read depth of greater than 10 reads and with FILTER = “PASS” by the xAtlas variant caller (8). The resulting proband VCFs were then merged using bcftools (v1.13) (9), and the variants were annotated via SpliceAI (v1.3.1) (masked scoring option) to determine variant effects on splice site acceptors or donors within 50bp of the variant (3). We filtered for those with delta SpliceAI scores of over 0.2, which is the lowest threshold to predict variants affecting splicing. The resulting variants were then annotated using Ensembl Variant Effect Predictor (VEP v102) (10). We prioritized variants with a gnomAD v3.1.1 (11) allele count of less than 20, a CADD (v1.6) score over 10 (12), SpliceAI gains and losses both scored over 0.2, and without an existing Pathogenic or Likely Pathogenic (P/LP) classification in ClinVar. Variants were then curated for those in genes associated with the respective proband cancer phenotype.

### RNA Analysis

Viably frozen peripheral blood mononuclear cells (PBMCs) from KCS probands were analyzed. PBMCs were thawed in a mixture of RPMI media and 10% fetal bovine serum (VWR, Radnor, PA) then allowed to incubate for 48 hours. Cells were incubated with or without 100μg/ml emetine (Sigma-Aldrich, St. Louis, MO), which inhibits translation and thereby prevents transcript degradation via nonsense mediated decay (NMD), for 6 hours. Total cellular RNA was extracted using Qiagen RNAeasy Micro kit (Qiagen, Chatsworth, CA). A two-step reverse transcriptase PCR (RT-PCR) analysis was performed: random hexamer primed Superscript first-strand synthesis system (Invitrogen, Waltham, MA) using 30ng of total RNA followed by PCR using two oligonucleotides (5’-TGACTTGCTATGTCGCCTTG-3’ and 5’-GGTCAGTTGCAGTTTCAGCA-3’). Products were gel purified using QIAquick Gel and PCR cleanup kit (Qiagen, Chatsworth, CA) and validated via Sanger sequencing (Eurofins Genomics, Louisville KY).

### Analysis of Variants Submitted to the ClinVar Database

Variants submitted to the ClinVar database (hosted by the National Center for Biotechnology Information) (13) within *DICER1* were acquired (October 2022). Intronic and synonymous variants with no predicted amino acid or termination change were annotated using SpliceAI (v1.3.1) and VEP (v102) as detailed in Methods - <u>Cohort Variant Analysis and Variant Filtration</u> (see above). Variants were prioritized for further analysis using the following criteria: 1) designated benign or likely benign in ClinVar, 2) absent from gnomAD v3.1.1 (11), and 3) both SpliceAI gains and losses scored over 0.2 and CADD (v1.6) score over 10 (12).

Variants submitted to ClinVar in *DICER1* and 60 other genes with Clinical Genome Resource (ClinGen) Variant Curation Expert Panel (VCEP) approved rules were downloaded (April 2023). Variants were filtered and annotated using the same methodology as above. For each variant tested via the *in vitro* splicing assay, the ClinVar submitters were contacted for phenotype information, when available.

### *In vitro* splicing assay

A splice reporter assay based on the vector system (pDESTSplice; AddGene #32484) was used following the protocol by Kishore, et al. (14). Desired sequences were either synthesized via gBlocks (IDT, Coralville, IA) or extracted from human genomic DNA (Promega, Madison, WI). The gBlock or extracted DNA harbored the exon-intron-exon junctions of interest along with 215 base pairs of intronic sequence, when applicable, flanking both exons and attB1 sites, created via primers for extracted DNA (Additional File 1: Table S2). Three different versions of each gBlocks were made: a reference version matching GRCh38, one with the allele of interest, and one with a common (per gnomAD database) non-reference allele near to the variants of interest (11) (Additional File 1: Table S2). Each gBlock was then cloned into pDONR221 using Gateway cloning following the manufacturer’s protocol (Invitrogen, Waltham, MA), verified by Sanger sequencing, and recombined into pDESTSplice. Extracted DNA was cloned into pDONR221 using Gateway cloning following the manufacturer’s protocol and the sequence was then verified. For regions not amenable to gBlocks, Phusion site-directed mutagenesis was performed following the manufacturer’s protocol (Thermo Scientific, Waltham, MA) using 5’-phosphorylated-primers (Additional File 1: Table S2) to generate the allele of interest or the common gnomAD allele in the pDONR221 vector containing reference sequence followed by sequence verification and recombination into pDESTSplice. Reporter clones were then isolated from bacteria using a QIAprep spin miniprep kit (Qiagen, Chatsworth, CA). Transfection of 800ng of each plasmid into 100k HEK-293T cells was done in triplicate using Lipofectamine 3000, following manufacturer’s protocol for 24-well plates (ThermoFisher, Waltham, MA). Following 24-hour incubation, total cellular RNA was extracted using Qiagen RNAeasy Micro kit. RT-PCR using 300ng of total cellular RNA was performed in two steps: random hexamer primed Superscript first-strand synthesis system (Invitrogen, Waltham, MA) followed by PCR using two oligonucleotides for rat insulin exons in pDESTSplice (5’-CCTGCTCATCCTCTGGGAGC-3’ and 5’-AGGTCTGAAGGTCACGGGCC-3’). Products were gel purified using QIAquick Gel and PCR cleanup kit (Qiagen, Chatsworth, CA) and analyzed on agarose gel electrophoresis and analyzed via Sanger sequencing (Azenta Life Sciences, South Plainfield, NJ). Intensity of gel bands was quantified using ImageJ 1.53t (15) and normalized to a background control. Graphs, simple linear regression, and statistics were generated using GraphPad Prism (v10).

## Results

### Texas KidsCanSeq Analysis

As a part of the CSER Consortium, the Texas KCS study recruited pediatric cancer probands at six clinical sites across Texas between 2018-2021 with solid tumors, lymphomas, or histiocytic disorders. Clinical germline exome and targeted panel sequencing was performed on 576 probands and reported in the medical record. Further analysis of the exome VCF files as described in Methods resulted in 242 variants in cancer predisposition genes of interest with both SpliceAI gains and losses scored over 0.2, absence in gnomAD v3.1.1, and CADD > 10. In total, three heterozygous variants in genes consistent with the patient’s tumor phenotype were identified. The first two variants were in *SUFU* (NM_016169.4:c.1177C>T(p.Arg393Trp)) in a medulloblastoma proband and in *RB1* (NM_000321.3:c.1960+1G>A) in a retinoblastoma proband. The former was reported as a variant of uncertain significance (VUS), and later determined to be generally inconsistent with proband’s cancer subtype, and the latter variant had been previously reported as pathogenic on the clinical exome and panel reports. Therefore, neither of these variants were selected for further functional study.

The third variant, NM_177438.3:c.574-26A>G in *DICER1* (termed Variant A) was from a patient with pleuropulmonary blastoma (PPB). The proband was diagnosed with PPB and was expected to have *DICER1-* related tumor predisposition syndrome given the parents also reported a family history of PPB in a paternal relative from another country with unclear work-up. However, no *DICER1* variants (pathogenic or uncertain) were on the clinical germline exome and germline panel reports. Variant A is intronic and 26 bp from the canonical splice acceptor at the 3’ end of the fourth intron of *DICER1*, outside the range of intronic variants typically evaluated by clinical platforms. Subsequent analysis of genomic DNA from either saliva (parents) or blood (proband) confirmed the presence of the intronic variant in the proband DNA and paternal transmission (Figure 1A). The splicing event predicted by SpliceAI is a loss of the canonical splice acceptor site and the creation of a splice acceptor site 20 bp upstream of the intronic variant, resulting in a 46 bp of inserted intronic sequence. Upon translation, this partial intronic retention would result in a PTC expected to trigger NMD of the *DICER1* mRNA (Figure 1B). RNA from viably frozen PBMCs from the PPB proband and another Texas KCS proband without PPB were analyzed by RT-PCR for products spanning exons 5 to 6 (Figure 1C). Sanger analysis confirmed the addition of a 46 bp insertion in 36% of the RNA from the PPB proband, with 64% reference sequence, and none from the control proband. The intronic variant may result in abnormal splicing due to disruption of the normal splicing branch point. As the proband’s PBMCs are heterozygous for the mutant allele (Figure 1D), these results suggest the majority of transcripts produced from the mutant haplotype result in the frameshift event.

**Figure 1.**
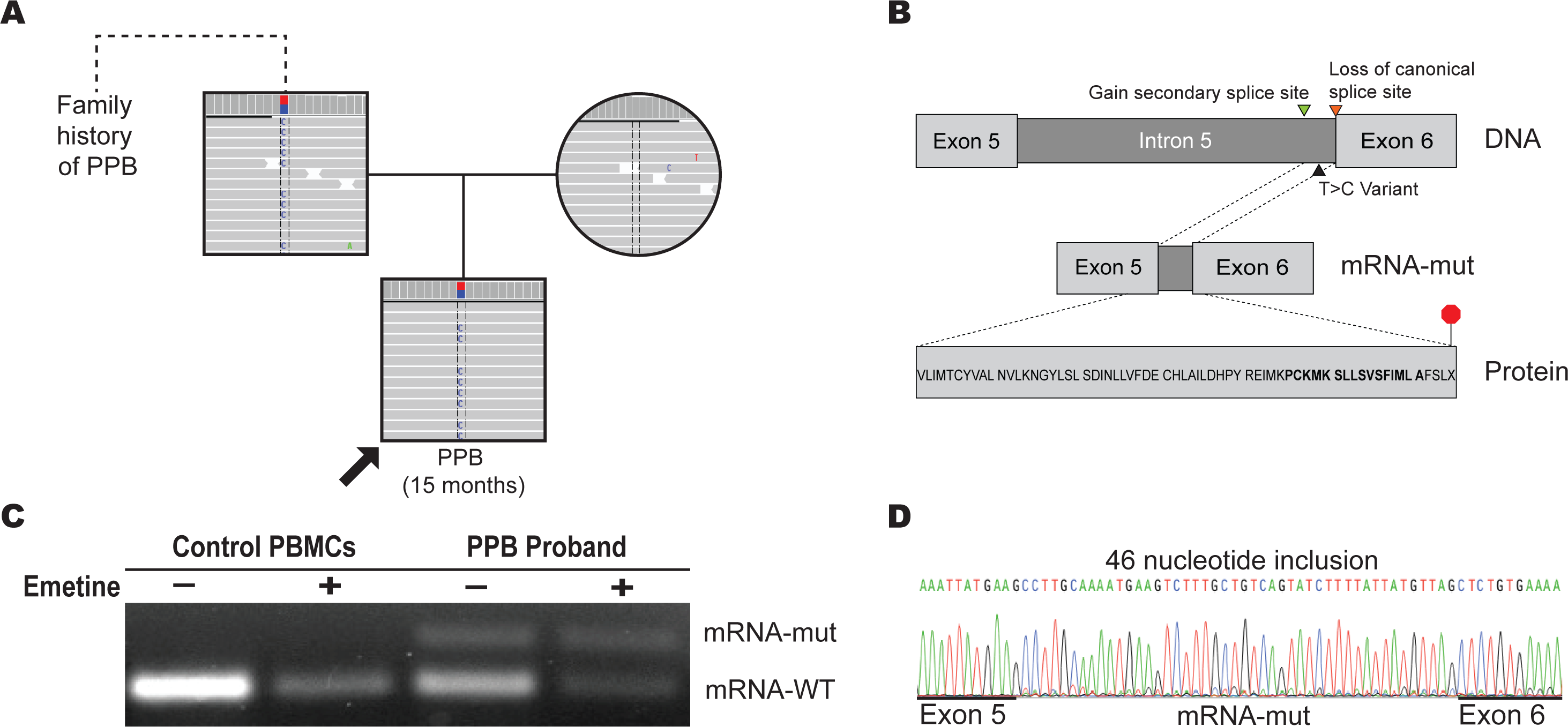
Intronic *DICER1* variant identified in KCS. **A** Pedigree showing the inheritance of said variant in the affected proband and how it fits with the family history of PPB. **B** Diagram outlining the predicted effect of the variant on splicing in the mRNA and how this change would impact translation, with the amino acids in bold coming from the inserted nucleotides. **C** Agarose gel image of the RT-PCR between exon 5 and 6 of *DICER1* with and without emetine of the PPB proband with the *DICER1* variant and a control patient without this variant. **D** Sanger sequencing showing the exon 5-6 junction of the mRNA-mut confirming the 46nt intron inclusion.

### ClinVar Variant Analysis – *DICER1*

Given the finding of this potentially clinically relevant *DICER1* variant in the KCS cohort, we searched for other variants designated as Benign and/or Likely Benign when submitted to ClinVar with similar predicted splicing abnormalities. To this end, all 4,227 *DICER1* variants in ClinVar were filtered using the methodology demonstrated in (Figure 2A) and described in Methods - <u>Analysis of Variants Submitted to the ClinVar Database</u>. This resulted in detection of a single synonymous variant, NM_177438.3(*DICER1*):c.5499G>A (Variant B) (Figure 3A, Table 1) that had been submitted to ClinVar once as Likely Benign. Inquiry of the submitting laboratory revealed that they had seen that variant in two unrelated probands: (1) an adult female with a history of recurrent thyroid cancer, parotid tail pleomorphic adenoma, and a parent with thyroid and colon cancer and (2) a child with a history of multinodular goiter, macrocephaly, and learning disability and a family history of thyroid nodules, goiters, thyroid cancer. Thus, both probands have phenotypes consistent with *DICER1*-related Tumor Predisposition Syndrome, which encompasses PPB, multinodular goiter, thyroid tumors and neurodevelopmental disorders (16). Neither of these probands were found to have any other pathogenic, likely pathogenic, or VUS results in *DICER1* or other genes tested at that time.

**Figure 2.**
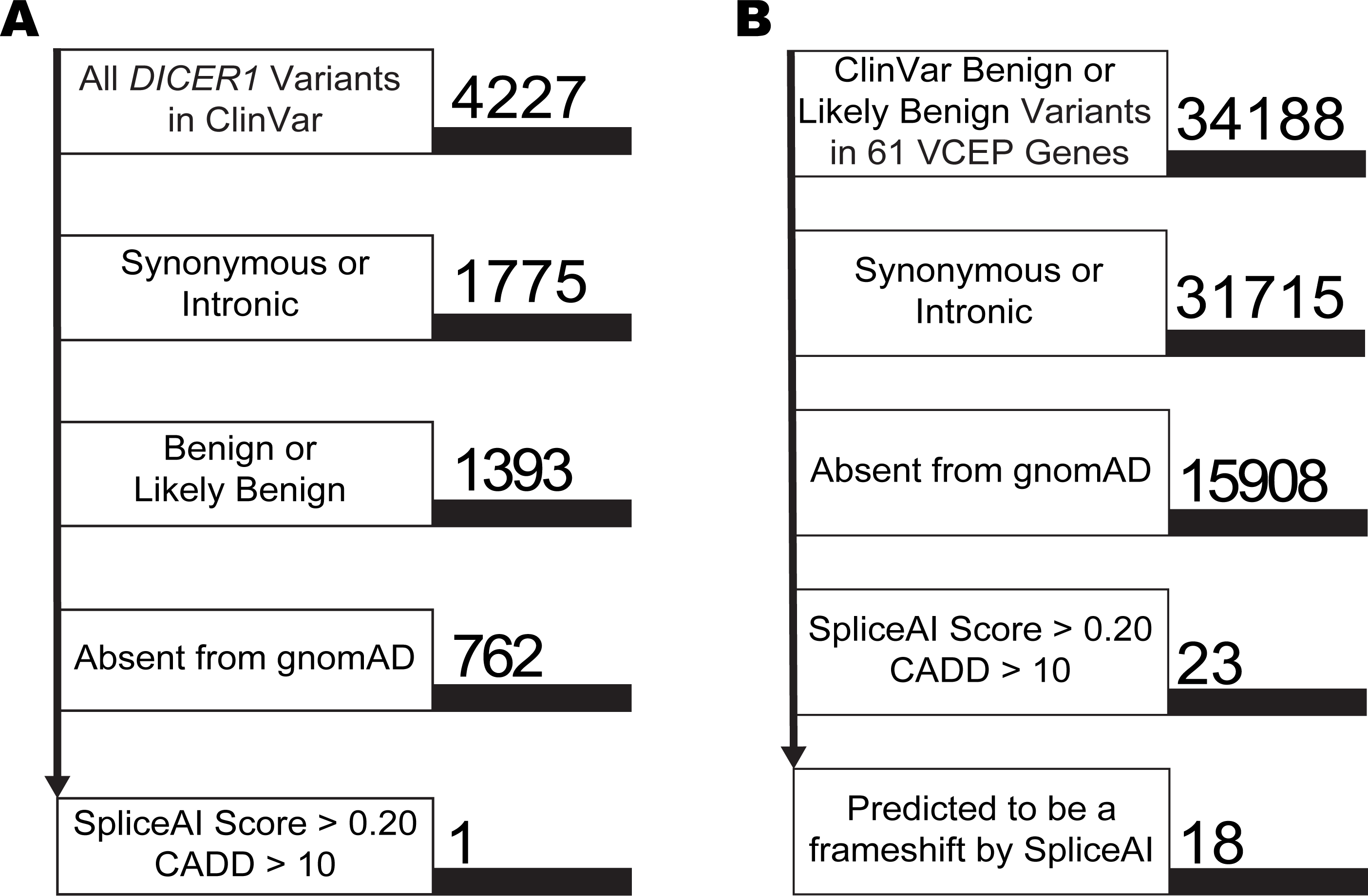
Identification of benign or likely benign variants in ClinVar predicted to missplice. **A** Prioritization of variants submitted to ClinVar in *DICER1* in initial analysis as of October, 2022. **B** Prioritization of variants submitted to ClinVar in 61 ClinGen Variant Curation Expert Panel (VCEP) genes, as of April, 2023.

**Figure 3.**
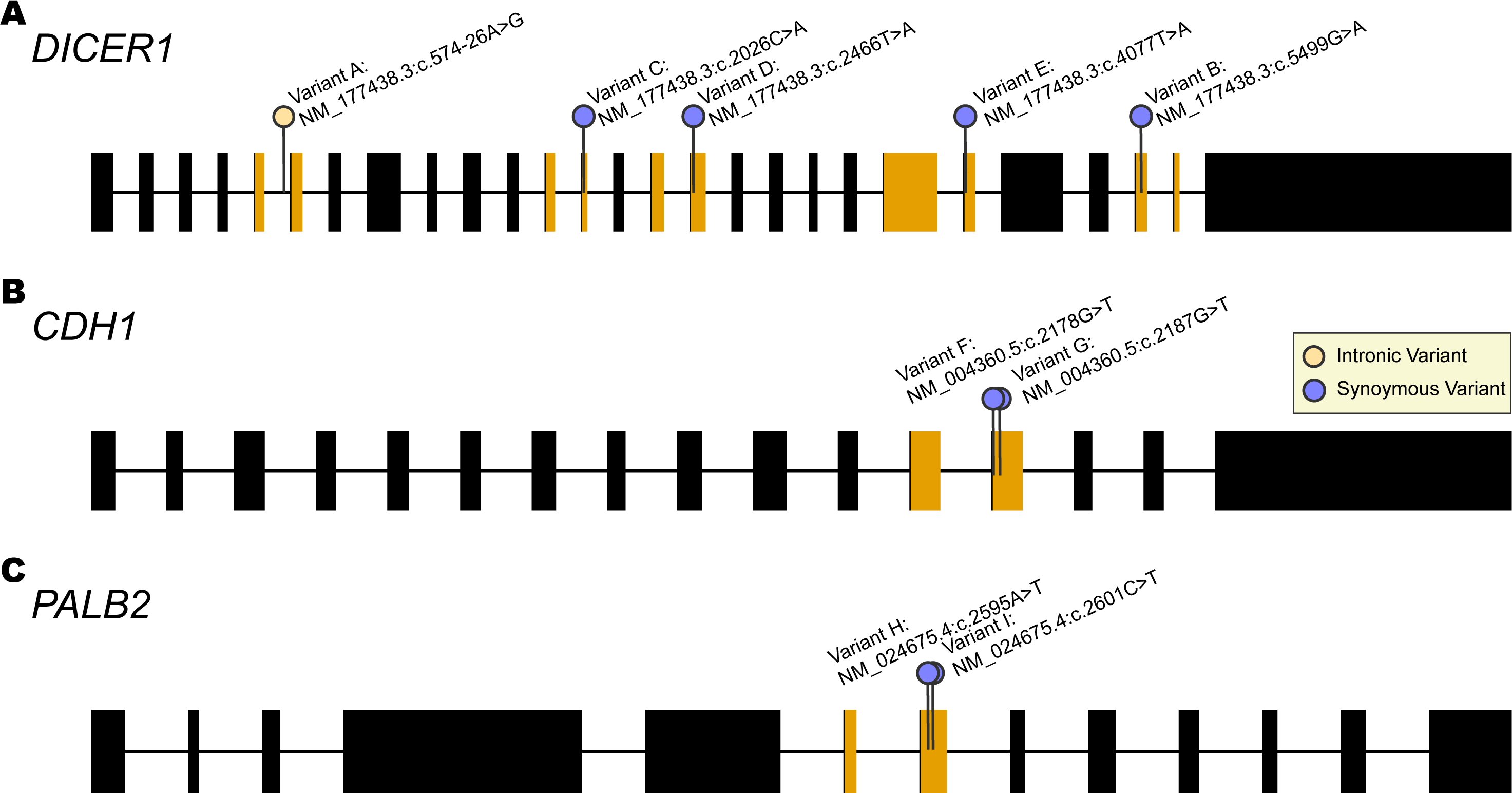
Location of assessed variants in **A** *DICER1*, **B** *CDH1*, and **C** *PALB2*. Colored exon-pairs indicates the exon-exon junctions used in each in vitro splicing assay.

**Table 1.**
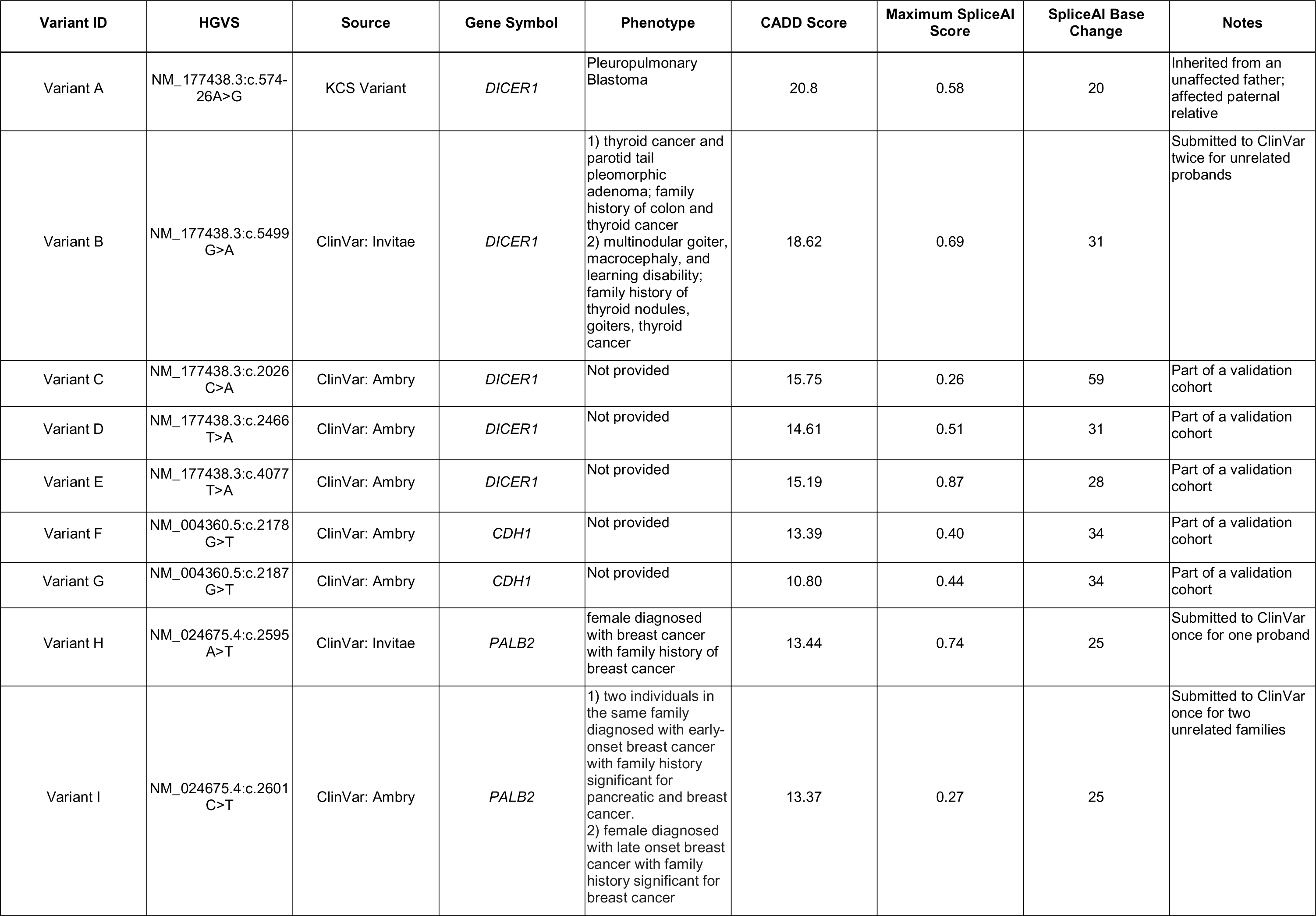
Overview of 9 variants assessed with CADD score, maximum SpliceAI delta score, and the relative position of predicted splice site loss or gain. Phenotype and notes provided when accessible.

| Variant ID | HGVS | Source | Gene Symbol | Phenotype | CADD Score | Maximum SpliceAI Score | SpliceAI Base Change | Notes |
| --- | --- | --- | --- | --- | --- | --- | --- | --- |
| Variant A | NM_177438.3:c.574-26A>G | KCS Variant | *DICER1* | Pleuropulmonary Blastoma | 20.8 | 0.58 | 20 | Inherited from an unaffected father; affected paternal relative |
| Variant B | NM_177438.3:c.5499G>A | ClinVar: Invitae | *DICER1* | 1) thyroid cancer and parotid tail pleomorphic adenoma; family history of colon and thyroid cancer<br>2) multinodular goiter, macrocephaly, and learning disability; family history of thyroid nodules, goiters, thyroid cancer | 18.62 | 0.69 | 31 | Submitted to ClinVar twice for unrelated probands |
| Variant C | NM_177438.3:c.2026C>A | ClinVar: Ambry | *DICER1* | Not provided | 15.75 | 0.26 | 59 | Part of a validation cohort |
| Variant D | NM_177438.3:c.2466T>A | ClinVar: Ambry | *DICER1* | Not provided | 14.61 | 0.51 | 31 | Part of a validation cohort |
| Variant E | NM_177438.3:c.4077T>A | ClinVar: Ambry | *DICER1* | Not provided | 15.19 | 0.87 | 28 | Part of a validation cohort |
| Variant F | NM_004360.5:c.2178G>T | ClinVar: Ambry | *CDH1* | Not provided | 13.39 | 0.40 | 34 | Part of a validation cohort |
| Variant G | NM_004360.5:c.2187G>T | ClinVar: Ambry | *CDH1* | Not provided | 10.80 | 0.44 | 34 | Part of a validation cohort |
| Variant H | NM_024675.4:c.2595A>T | ClinVar: Invitae | *PALB2* | female diagnosed with breast cancer with family history of breast cancer | 13.44 | 0.74 | 25 | Submitted to ClinVar once for one proband |
| Variant I | NM_024675.4:c.2601C>T | ClinVar: Ambry | *PALB2* | 1) two individuals in the same family diagnosed with early-onset breast cancer with family history significant for pancreatic and breast cancer.<br>2) female diagnosed with late onset breast cancer with family history significant for breast cancer | 13.37 | 0.27 | 25 | Submitted to ClinVar once for two unrelated families |

### *In vitro* splice reporter assay

Biological samples were not available from either patient with the *DICER1* ClinVar variant. We thus decided to evaluate the KCS variant and this ClinVar variant with a well-established *in vitro* splice reporter assay. The reporter vector contains integration of the *DICER1* exon-intron-exon sequence between two rat insulin exons that are driven by the Rous sarcoma virus long terminal repeat promoter. We designed fragments encompassing the reference allele for the relevant section of *DICER1*, the mutant alleles identified in KCS (Variant A) or ClinVar (Variant B) variant and a common variant identified in gnomAD as a control (Figure 4A; Additional File 1: Table S2). After transfection into HEK-293T cells, RNA products are quantified and sequenced to assess the effects of variation on splicing efficiency and accuracy. As shown in figure 4B, the RNA produced for the reference vector and gnomAD-common variant vector were consistent with the MANE Select transcript. The Variant A splicing assay recapitulated the abnormal splice products seen in the monocyte-derived RNA of the Texas KCS proband (Figure 1D) at 52% of splicing products; in addition, two secondary mRNAs were detected, at 30% and 18% abundance, that were not observed in the proband analysis; both of these products also result in a frameshift (Figure 4C). Results from Variant B analysis identified the SpliceAI predicted 31-nucleotide exclusion of the 3’-end of exon 25 (100%) compared with normal products for the reference and gnomAD variant vector (Table 1; Figure 5A). The resulting 31-nucleotide frameshift mRNA would be expected to undergo NMD; even if truncated protein were produced, the critical *DICER1* double-stranded RNA binding domain would be disrupted. Of note, this variant has been subsequently upgraded to VUS by the submitting lab given the SpliceAI score.

**Figure 4.**
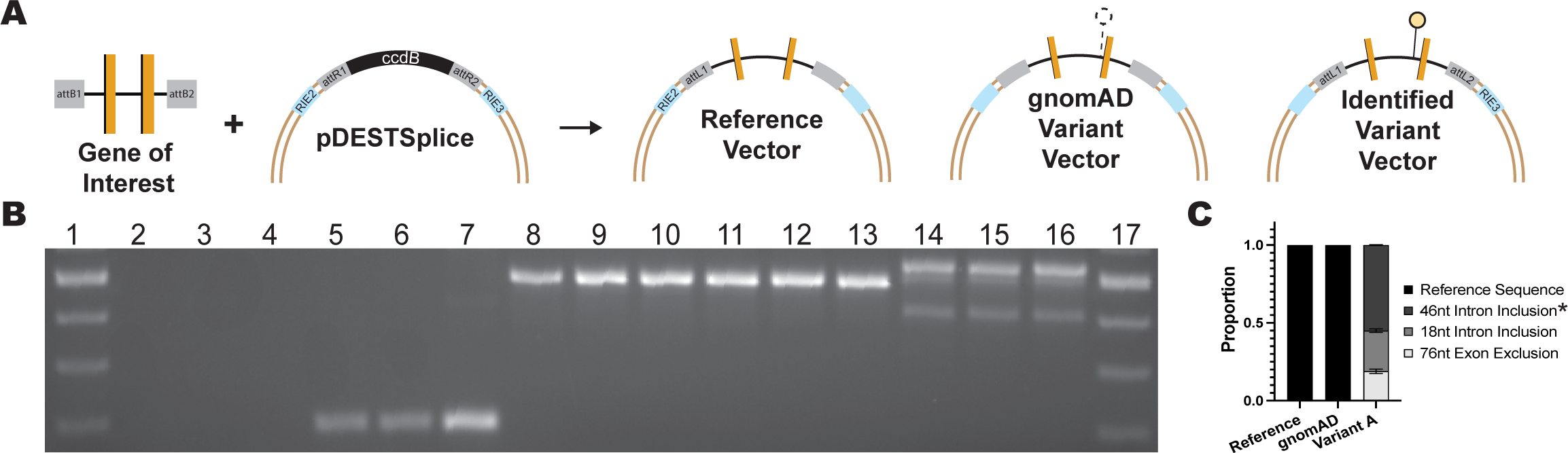
Diagram and result of variant identified in KCS using in vitro splicing assay. **A** Diagram of the vectors generated from the Gene of Interest using pDESTSplice, with each vector containing either the reference, a common gnomAD variant, or the identified variant of interest. **B** Results of the RT-PCR of Variant A, identified in KCS, where lanes 1 and 17 are size markers, 2-4 are non-transfected controls, 5-7 are empty pDESTSplice, 8-10, is the reference vector,11-13 is the vector containing the common gnomAD variant, and 14-16 are the vector containing the identified variant of interest. **C** Graph showing the proportion of products produced by RT-PCR in the reference and variant vector, identifying the proportion of products produced through quantification of luminescence, with the * denoting the mRNA-mut identified in the patient’s RNA, as shown in Figure 1C.

**Figure 5.**
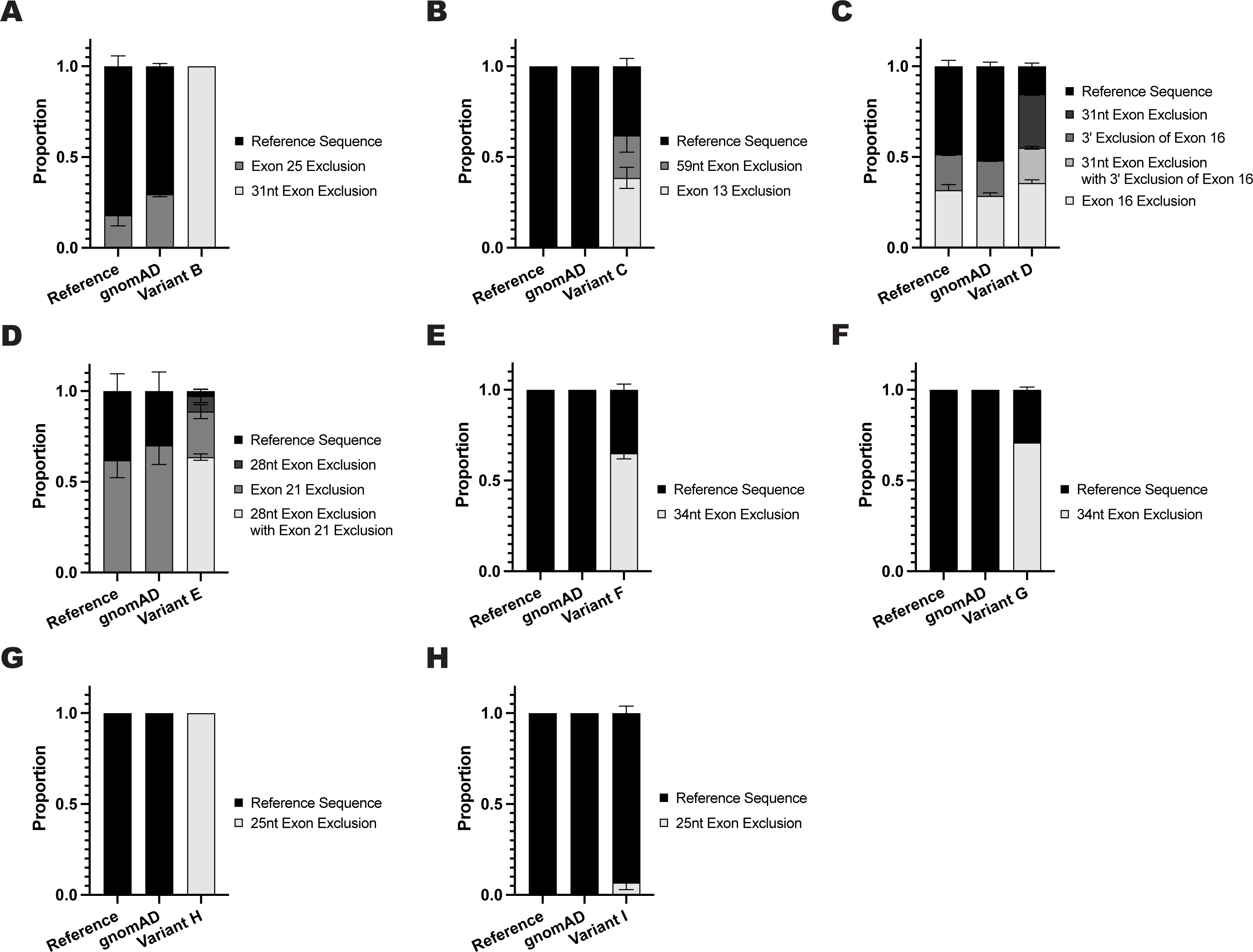
Result of in vitro splicing assay on Benign/Likely Benign variants extracted from ClinVar. Panel **A-H** shows results of RT-PCR for Variant B through Variant I displaying the proportion of products produced by RT-PCR in the reference, nearby common variant, and variant vector. Photographs of the original gel electrophoresis are available in Additional File 2: Figure S4.

### ClinVar Variant Analysis - 61 ClinGen Expert Panel Genes

Given the results of the *in vitro* splice reporter assay, we expanded the ClinVar analysis to all 61 genes with Clinical Genome Resource (ClinGen) variant curation expert panel (VCEP) specifications, which includes *DICER1*. We selected ClinGen VCEP genes, as ClinGen has recently published recommendations for incorporation of SpliceAI into variant classification specifications (17). We extracted 34,188 Benign/Likely Benign variants across the 61 genes that were subsequently filtered using the methodology described in Methods - <u>Analysis of Variants Submitted to the ClinVar Database</u> (Figure 2B). Among those, 23 variants had both SpliceAI gains and losses with scores over 0.2, CADD scores > 10, and were absent from gnomAD (Figure 2B). Eighteen of these variants, affecting 12 genes, are predicted to lead to a frameshift when abnormal splicing occurs and should be considered for additional pathogenicity analysis (Additional File 1: Table S4). Four of these variants occurred in *DICER1*, including Variant B and three submitted after our initial analysis (Table 1).

To further assess evidence for missplicing, we performed *in vitro* splicing assays on the new *DICER1* variants, two variants in *CDH1* (Table 1; Figure 2B) and two in *PALB2* (Table 1; Figure 2C). These variants were selected because of their potential clinical importance and for each of these variants, the full-length exon and intron sequences flanking the variant could be incorporated into the splicing vector. As previously, a nearby common gnomAD variant was selected as a control (Additional File 1: Table S3). As shown in figure 5 panels B-H, the *in vitro* splice products for both the reference and gnomAD common alleles were similar, while the prioritized mutant alleles all revealed abnormal splice products not present in the controls to varying extent ranging from 6.8% (Variant I) to 100% (Variant H) (Additional File 2: Figure S1A). Of the abnormal products, the product predicted by SpliceAI was the largest component in all but two variants (Variant D at 49.2% and Variant I at 6.8%), however, the other abnormal products would also lead to frameshifts and subsequent PTCs (Figure 5 B-H; Additional File 2: Figure S2).

We analyzed the SpliceAI scores for predicted loss and gain versus the proportion of mutant products and see some correlation consistent with the recent ClinGen guidance to use 0.2 as a minimum SpliceAI cut-off which would need validation in a larger study. Comparing mean percentage of abnormal product produced versus SpliceAI scores, the strongest correlation appears to be the SpliceAI donor or acceptor loss score which could be studied further in a larger series (Additional File 2: Figure S1).

After completing these assays, we reached out to the submitting laboratories for any clinical information. Five of these variants (Variants C-G Table 1) were part of a validation cohort without any clinical information. Variant H in *PALB2* was found in a female proband with breast cancer with a family history of breast cancer. Variant I, also in *PALB2*, was found to occur in three patients from two families. In the first family, two individuals carried the variant and were diagnosed with breast cancer, and there is an additional family history significant for pancreatic cancer and breast cancer (including a male relative). The third unrelated patient was a female diagnosed with late onset breast cancer with family history significant for breast cancer (also in a male relative). There were no other reportable variants identified for any of the three kindreds. The cancer diagnoses in these three kindreds are consistent with the phenotype of individuals with monoallelic *PALB2* loss-of-function variants (18, 19) as well as monoallelic *BRCA1* or *BRCA2* loss-of-function variants.

### Application of ACMG Classification guidelines

We applied the *DICER1* VCEP specifications (clinicalgenome.org, Accessed 19 January 2023) for the two *DICER1* variants (A and B) where clinical phenotype information was available. The following evidence codes were applied: 1. PS4_Supporting: the variant is absent from controls and the gene is strongly associated with the proband disease, PPB; 2. PM2_Supporting: the variant is absent from the gnomAD v 3.1.1 database. When using the PP3 code for the application of *in silico* tools, initially the *DICER1* VCEP recommended that Supporting Pathogenicity weight required “[f]or splicing variants, concordance of MaxEntScan and SpliceAI” (clinicalgenome.org, Accessed 19 January 2023). While SpliceAI predicts both variants to influence splicing, these predictions could not be corroborated with evidence from MaxEntScan (4, 20) as neither variant qualifies for MaxEntScan scores given their distance from the canonical splice site. Recently, updated guidelines from the *DICER1* VCEP, have clarified the BP4 and PP3 application for variants outside of the MaxEntScan window to only require SpliceAI prediction. Incorporation of the *in vitro* splice reporter assay would add the PS3 evidence code, for application of functional assays, given both variants result in an experimentally confirmed out-of-frame impact on splicing. Together this could result in designation of Likely Pathogenic to both variants.

## Discussion

We have identified potentially clinically significant variants originally classified as Likely Benign or in intronic regions not usually considered within reportable range in patients with phenotypes representative of *DICER1*-related Tumor Predisposition Syndrome. The approach described here that incorporates allelic frequency, SpliceAI prediction, and CADD score identified these variants in KCS exome data and the ClinVar database across many genes with minimal analytical labor and high specificity. Incorporation of this information can result in clinically meaningful changes in variant classification. For example, Variant A became a clear candidate for biological validation given that 1) there is a strong connection between the *DICER1* variant and the proband’s and his relatives phenotype, 2) SpliceAI predicted both an acceptor loss and gain above 0.20, and 3) no other pathogenic variant in *DICER1* had been reported. Functional assessment results in the designation of Variant A as Likely Pathogenic.

We focused on intronic and synonymous Likely Benign variants because they are not typically reported to the requesting physician and thus may not be further analyzed for pathogenicity. Interestingly, analysis of ClinVar for nonsynonymous Likely Benign variants returned no variants that met gnomAD allele frequency, CADD, and SpliceAI filtering criteria. Of note, although not analyzed further, there are many variants currently classified as VUSs in ClinVar that meet these criteria and have the potential to be pathogenic or likely pathogenic pending further assessment.

Potential misclassification of disease-relevant variants as B/LB may represent a systematic problem with the sole use of older algorithms designed to detect potential splicing defects. Variants that are farther from the splice site, either intronic or synonymous, are typically not subject to functional evaluation by clinical pipelines. As seen in Variant B (2 families), Variant H (1 family), and Variant I (2 families), patients with no other reportable variants displayed phenotypes consistent with monoallelic loss-of-function variants, in *DICER1* and *PALB2* respectively. It is important to note that all 9 of the variants predicted by SpliceAI to cause missplicing result in the use of aberrant splice sites (at distances up to dozens of nucleotides away) that are not found in any known transcript and are outside the range of other splice prediction algorithms (4, 21). Biological validation through analysis of patient RNA and *in vitro* splicing assays confirm: 1) the limitations of only using a scoring algorithm that does not characterize all variants; 2) the scoring guidelines for SpliceAI from the original paper [“confidently predicted cryptic splice variants(score ≥ 0.5)”] are too conservative; and 3) variants scoring above 0.20 via SpliceAI should be considered potential missplicing variants consistent with the recently published ClinGen guidance (3, 17). Across the nine variants assessed, we also identified a correlation between increased SpliceAI scores and abnormal products produced through the *in vitro* splicing assay, with the strongest correlation being seen with the increased SpliceAI donor or acceptor loss score (Additional File 2: Figure S1C). Though this is a limited analysis, splice variants do not always affect splicing in a binary manner, and it is possible that increasingly large SpliceAI scores may act as further evidence for classifying a variant’s effect on splicing (i.e., while 0.2 is a useful threshold, larger values may provide even more evidence) as seen for missense predictors (22). We have additionally demonstrated that as SpliceAI donor or acceptor loss scores decrease towards the 0.2 cut-off, the proportion of abnormal products detected by our splice reporter assays are increasingly variable. This phenomenon may not reflect the true biological consequences of these variants in patient transcriptomes.

The implementation of newer tools such as SpliceAI, demonstrates the ongoing improvement of algorithms and meta-predictors in describing potential effects of variants to shorten diagnostic odysseys for patients. Although large-scale, massively parallel splicing assays are beginning to be available to support these predictions (23, 24), the combined use of clinical RNA and DNA analysis can also reveal abnormal splicing products and aid in the rapid identification of clinically relevant variants (25, 26). Laboratories should also consider re-evaluation of previously classified variants with these newer algorithms to consider variant reclassification given the importance of diagnoses on proband outcomes and at-risk relatives obtaining prevention measures.

## Conclusions

We leveraged SpliceAI to identify synonymous and intronic variants that alter splicing and were either overlooked or originally classified as Likely Benign. Using an *in vitro* splice reporter assay, we demonstrated that the variants identified using this methodology across multiple different genes lead to the predicted abnormal splice product to a varying extent and should be considered for reclassification towards pathogenicity. These variants illustrate the potential for action at a distance from canonical splice sites on altering splice products and the relevance of these effects to rare disease. Our results also underscore the benefits of combined DNA and RNA studies in evaluating previously molecularly undiagnosed patients.

## Data Availability

Genome sequencing and phenotype data for study participants that opted-in to data sharing are available for authorized access and hosted via dbGaP for the Texas KCS study (phs002378.v1.p1). We have submitted the individual variants found to be of potential clinical relevance to ClinVar; accession IDs for each variant described can be found in revised Additional File 1: Table S4.

## Abbreviations

B/LB: Benign or Likely Benign
ACMG/AMP: American College of Medical Genetics and Genomics/Association for Molecular Pathology
CADD: Combined Annotation Dependent Depletion
PTC: Premature Termination Codon
KCS: KidsCanSeq Study
CSER: Clinical Sequencing Evidence-Generating Consortium
VCF: Variant Call File
VEP: Variant Effect Predictor
P/LP: Pathogenic or Likely Pathogenic
PBMC: Peripheral Blood Mononuclear Cells
NMD: Nonsense Mediated Decay
RT-PCR: Reverse Transcriptase PCR
ClinGen: Clinical Genome Resource
VCEP: Variant Curation Expert Panel
VUS: Variant of Uncertain Significance
PPB: Pleuropulmonary Blastoma
DCIS: Ductal Carcinoma In Situ

## DECLARATIONS

### Ethics approval and consent to participate

The KCS study was approved by the Institutional Review Board (IRB) of Baylor College of Medicine which served as the central IRB for all participating sites (protocol number H-42376).

### Authors Contributions

Development of KCS Protocol: SEP, DWP. Enrollment and clinical description of KCS participant: SPR, KLV. Identification of splicing variants in KCS: SAF. KCS patient RNA analysis: ORH. Inheritance analysis of KCS PPB patient: DWP, AR. Identification of B/LB variants in ClinVar: SAF, ORH. *In vitro* splicing assays and analysis: ORH. Writing—original draft: SAF, ORH. Writing—review and editing: SAF, ORH, DWP, AR, SEP, GMC. Resources: SEP and DWP. All authors read and approved the final manuscript.

## Acknowledgments

The authors would like to acknowledge Ambry Genetics and Invitae for providing the phenotype information included in the manuscript. We also acknowledge the families that agreed to participate in KCS Study.

## SUPPLEMENTARY INFORMATION

### Additional File 1

This additional file consists of the following supplementary tables referenced throughout the manuscript: **Table S1.** List of 181 cancer predisposition genes screened in analysis of Texas KCS proband exomes. **Table S2.**

List of oligonucleotides used in in vitro splice assay. **Table S3.** List of nearby benign variants identified in gnomAD and used in in vitro splice assay. **Table S4.** Overview of 23 variants resulting from the SpliceAI screen of variants submitted to the ClinVar Database with CADD score, maximum SpliceAI delta score, and the relative position of predicted splice site loss or gain.

### Additional File 2

This additional file consists of the following supplementary figures referenced throughout the manuscript: **Figure S1.** Graphs of mean percentage of abnormal and normal products produced in respective in vitro splicing assay. **Figure S2.** Diagram of all products produced in mini-gene assays order as seen on gels, corresponding to Variants A-I. **Figure S3.** Gel electrophoresis images of RT-PCR, corresponding to variant B-I.

