## Additional File 2 for "Combined Bioinformatic and Splicing Analysis of Likely Benign Intronic and Synonymous Variants Reveals Evidence for Pathogenicity"

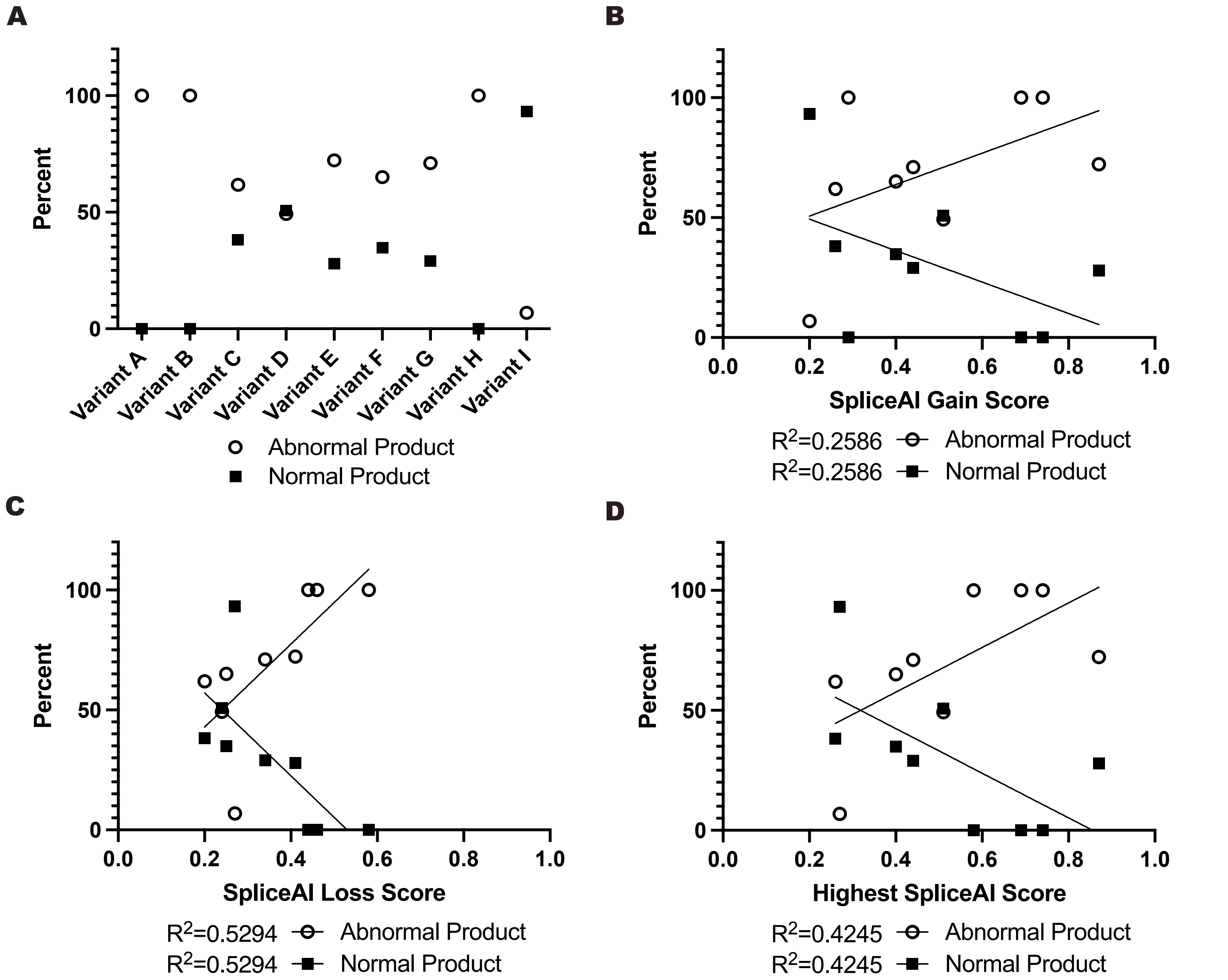


**Figure S1.** Graphs of mean percentage of abnormal and normal products produced in respective in vitro splicing assay. **A** Per variant proportion of products produced across all nine variants assessed. **B** Correlation between SpliceAI score of gained secondary splice site and mean percentage of abnormal and normal products with corresponding $R^2$ (F-test, *P*-value = 0.1621). **C** Correlation between SpliceAI score of lost primary splice site and mean percentage of abnormal and normal products with corresponding $R^2$ (F-test, *P*-value = 0.0263). **D** The correlation between the variant's highest SpliceAI score and mean percentage of abnormal and normal products with corresponding $R^2$ (F-test, *P*-value = 0.0573).

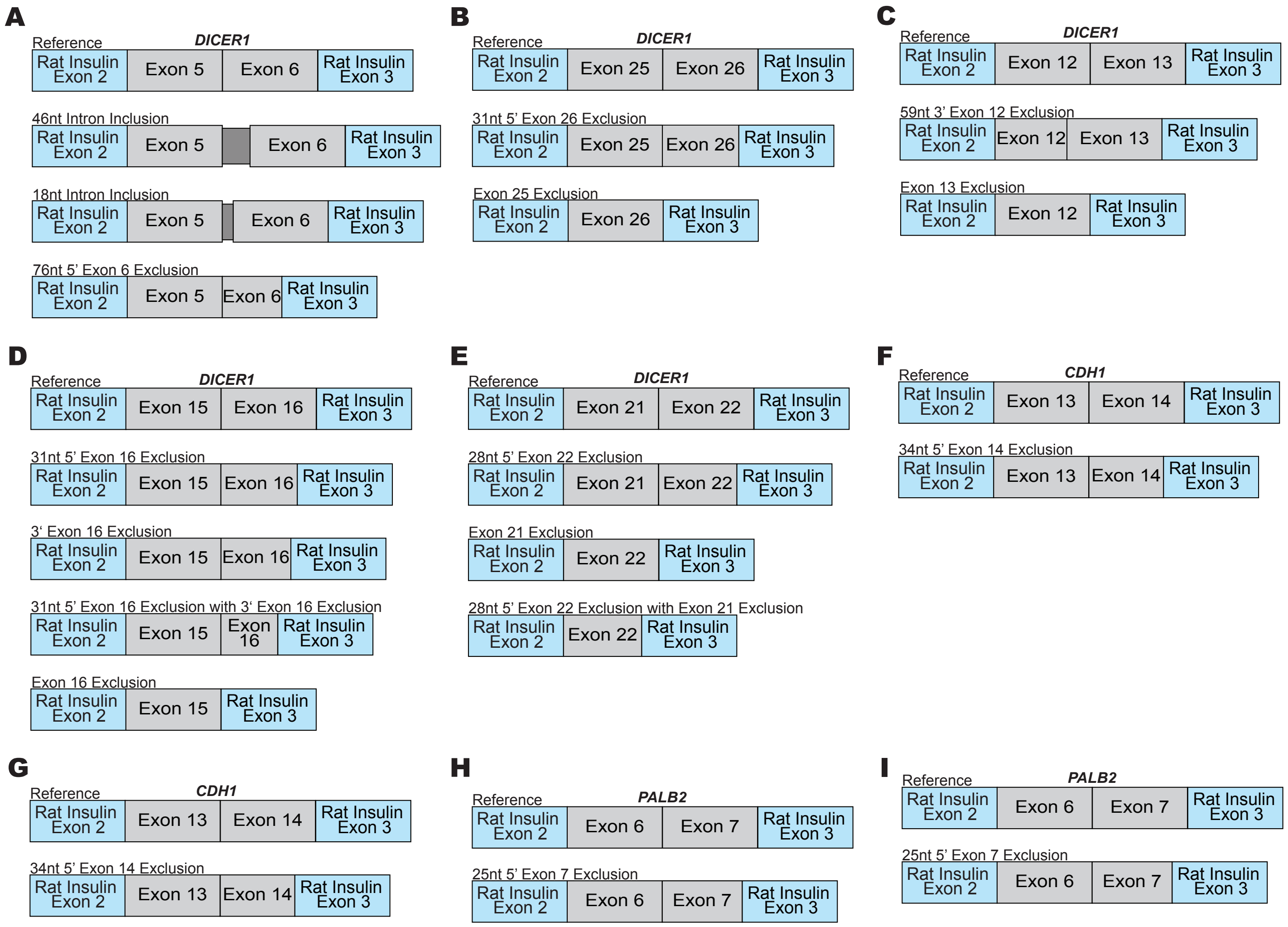


**Figure S2. A-I** Diagram of all products produced in mini-gene assays order as seen on gels, corresponding to Variants A-I.

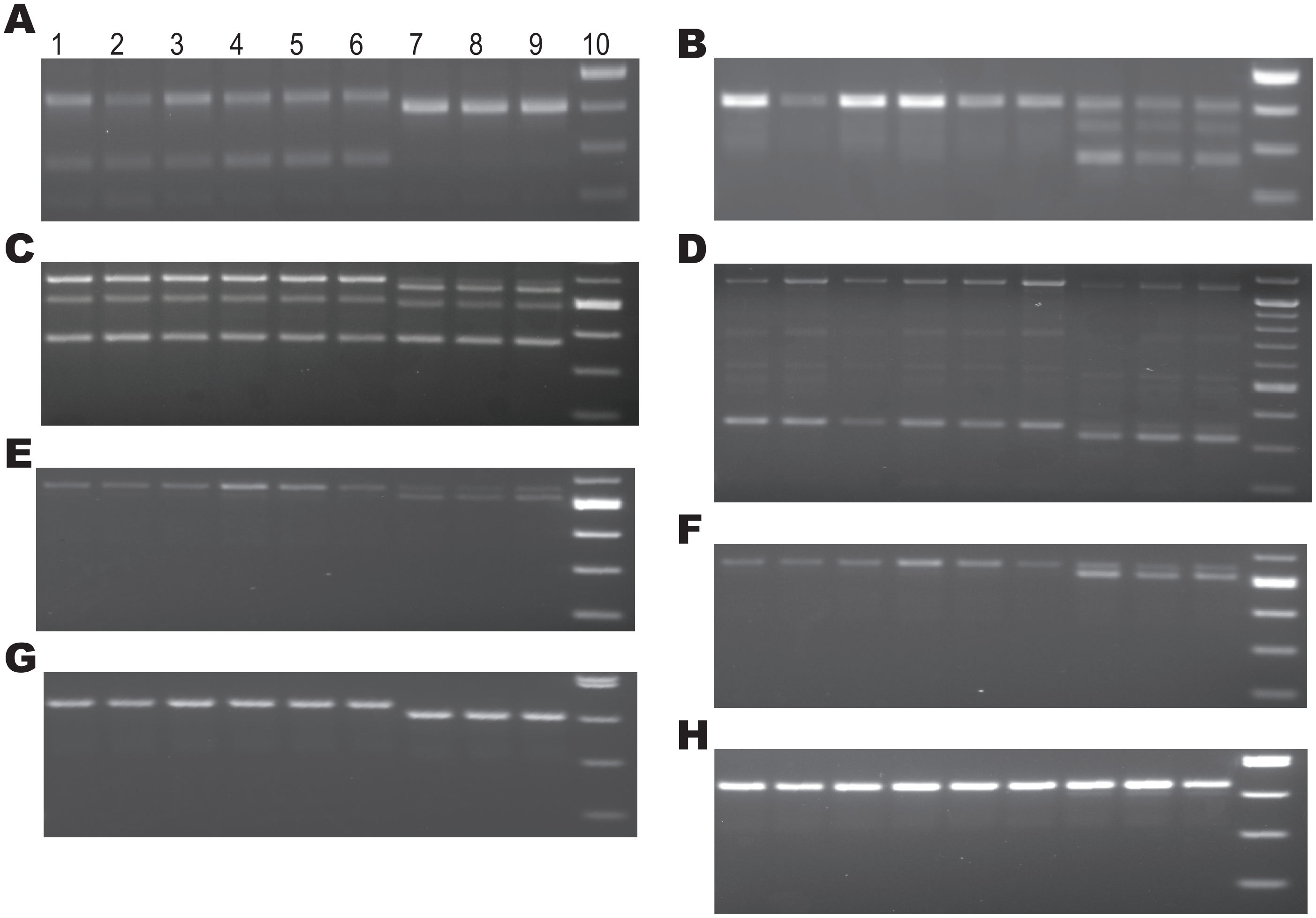


**Figure S3. A-H** Gel electrophoresis images of RT-PCR, corresponding to variant B-I, where 1-3, is the reference vector, 4-6 is the vector containing the common gnomAD variant, and 7-9 are the vector containing the identified variant of interest, where 10 is a size marker.
